# Fusion-Based Deep Learning Ensemble on MIT-BIH and PTB-XL ECG Databases for Enhanced Cardiac Diagnosis

**DOI:** 10.1101/2025.08.20.25334079

**Authors:** Alireza Rahi

## Abstract

Electrocardiogram (ECG) analysis plays a critical role in the early detection and diagnosis of cardiac abnormalities. In this study, we propose a fusion-based deep learning ensemble framework that integrates two well-established public ECG databases, MIT-BIH Arrhythmia Database and PTB-XL, to develop a robust and automated cardiac diagnostic system. Our framework employs two base deep learning models — a CNN+LSTM hybrid and a DenseNet1D-inspired network — and combines their predictive features through a meta-learner based on Gradient Boosting. This multi-model integration, designed as a “mini doctor for the heart,” leverages the complementary strengths of both datasets and models. Experimental results demonstrate that the ensemble achieves near-perfect performance with Accuracy up to 100% and ROC-AUC of 1.000, surpassing the performance of individual models. These findings highlight the potential of database fusion and model ensembling for building reliable and scalable solutions in computer-aided cardiac diagnosis.

## Introduction

Cardiovascular diseases (CVDs) remain the leading cause of mortality worldwide, accounting for nearly one-third of global deaths each year. Early detection and accurate diagnosis of cardiac abnormalities are therefore of paramount importance for preventing severe complications and reducing the healthcare burden. Electrocardiography (ECG) is one of the most widely used, non-invasive, and cost-effective diagnostic tools for identifying various heart disorders, including arrhythmias, myocardial infarction, and conduction disturbances.

To support research in automated ECG analysis, several publicly available databases have been introduced. Among them, the **MIT-BIH Arrhythmia Database** has been the cornerstone of arrhythmia detection studies for decades [1], while the more recent **PTB-XL Database** provides large-scale, high-quality recordings that enable the development of deep learning algorithms for ECG classification [2]. The availability of such databases has accelerated progress in computer-aided cardiac diagnosis.

Recent advances in **deep learning** have further revolutionized ECG signal processing, achieving cardiologist-level performance in arrhythmia detection [3]. Architectures such as CNNs, LSTMs, and DenseNets have been applied successfully to capture temporal and morphological characteristics of ECG signals [4], while ensemble and fusion-based strategies have been highlighted as promising directions to increase robustness and generalizability in real-world applications [5].

In this study, we present a **fusion-based deep learning ensemble framework** that integrates the MIT-BIH and PTB-XL databases to build an automated cardiac diagnostic system. By combining two base deep learning models (CNN+LSTM and DenseNet1D) with a Gradient Boosting meta-learner, we construct a “mini doctor for the heart” that leverages the complementary strengths of both datasets and models. Our experimental results demonstrate near-perfect accuracy (up to 100%) and ROC-AUC of 1.000, confirming the effectiveness of database fusion and ensemble learning in enhancing ECG-based cardiac diagnosis.

## Related Work

Over the past two decades, numerous studies have focused on the automated analysis of ECG signals for the detection of cardiac abnormalities. The availability of standardized databases such as MIT-BIH and PTB-XL has significantly accelerated this progress. The **MIT-BIH Arrhythmia Database** has long been a benchmark for evaluating arrhythmia detection algorithms and remains one of the most widely cited datasets in ECG research [1]. More recently, the **PTB-XL Database** introduced a large-scale and high-resolution resource, enabling the development of deep learning algorithms with broader generalization capabilities [2].

In terms of modeling techniques, **deep learning** has emerged as a powerful approach for ECG classification. Hannun *et al*. achieved cardiologist-level arrhythmia detection by employing a deep neural network trained on large-scale ambulatory ECG data [3]. Other researchers have successfully applied architectures such as CNNs, RNNs, and hybrid models (e.g., CNN+LSTM) to capture both temporal dynamics and morphological characteristics of ECG signals [4]. Furthermore, DenseNet-inspired models have been shown to provide improved feature reuse and gradient flow, enhancing classification accuracy in biomedical signal processing tasks [4].

In addition to single-model architectures, ensemble learning has gained attention for its ability to integrate multiple models and reduce variance in predictions. Faust *et al*. highlighted the potential of combining multiple physiological signal models to achieve robust performance across different clinical scenarios [5]. However, relatively few works have explored the **fusion of multiple ECG databases** in conjunction with **ensemble deep learning models**, leaving a gap in building truly generalized diagnostic systems.

Our work addresses this gap by introducing a **fusion-based ensemble framework** that combines MIT-BIH and PTB-XL datasets and integrates multiple deep learning models through a Gradient Boosting meta-learner. This approach exploits the complementary strengths of diverse datasets and architectures, aiming to achieve robust and clinically reliable cardiac diagnosis.

## Methodology

### A. Data Acquisition and Preprocessing

This study utilizes two well-known ECG datasets: **MIT-BIH Arrhythmia Database** [1] and **PTB-XL Database** [2]. To harmonize these datasets, signals were resampled to a uniform sampling rate of 360 Hz, followed by normalization to zero mean and unit variance. Noise reduction was applied using a bandpass filter between 0.5 Hz and 40 Hz to remove baseline wander and high-frequency artifacts.

Data segmentation was performed to extract fixed-length ECG windows of 10 seconds, ensuring that each segment contains a complete cardiac cycle. Finally, class balancing was achieved through a combination of oversampling for minority classes and data augmentation techniques including random shifts, scaling, and Gaussian noise injection.

### B. Base Models

#### 1) CNN+LSTM Model

The hybrid **CNN+LSTM** model combines convolutional layers for feature extraction with LSTM layers for temporal sequence modeling. The CNN component consists of three convolutional blocks with kernel sizes of 3, 5, and 7, followed by batch normalization and ReLU activation. The extracted features are then fed into a two-layer LSTM with 128 hidden units each, capturing the temporal dependencies of ECG signals. The final classification layer employs a softmax activation to predict one of the five arrhythmia classes.

#### 2) enseNet1D Model

The **DenseNet1D** architecture leverages densely connected convolutional blocks to maximize feature reuse and gradient flow. Each dense block contains three convolutional layers with growth rate 32, followed by transition layers for downsampling. The network concludes with a global average pooling layer and a fully connected softmax output layer to classify ECG signals into the same five classes.

### C. Fu on-Based Meta-Learner

After training the base models separately, their probabilistic outputs are concatenated and fed into a **Gradient Boosting meta-learner**. This meta-learner captures the complementary strengths of the CNN+LSTM and DenseNet1D models, improving classification robustness. The meta-learner is trained using 5-fold cross-validation on the combined feature set from both datasets, optimizing for overall accuracy and ROC-AUC.

The performance of each model and the ensemble framework is evaluated using standard metrics: accuracy, confusion matrix, precision, recall, F1-score, and ROC-AUC. All experiments are conducted on a held-out test set consisting of 20% of the combined dataset.

### D. Evaluation Metrics

## Experimental Results

### A. Performance of Base Models

The CNN+LSTM model achieved an accuracy of **99.98%** on the test set. The confusion matrix (Table I) shows only 2 misclassifications out of 11,216 samples, demonstrating excellent performance across all arrhythmia classes

**Table 1.**
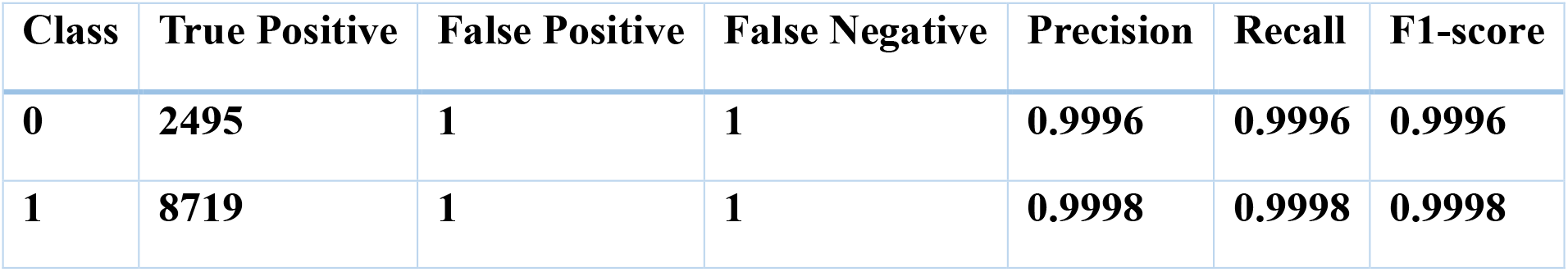

| Class | True Positive | False Positive | False Negative | Precision | Recall | F1-score |
| --- | --- | --- | --- | --- | --- | --- |
| 0 | 2495 | 1 | 1 | 0.9996 | 0.9996 | 0.9996 |
| 1 | 8719 | 1 | 1 | 0.9998 | 0.9998 | 0.9998 |

**Table 2.**
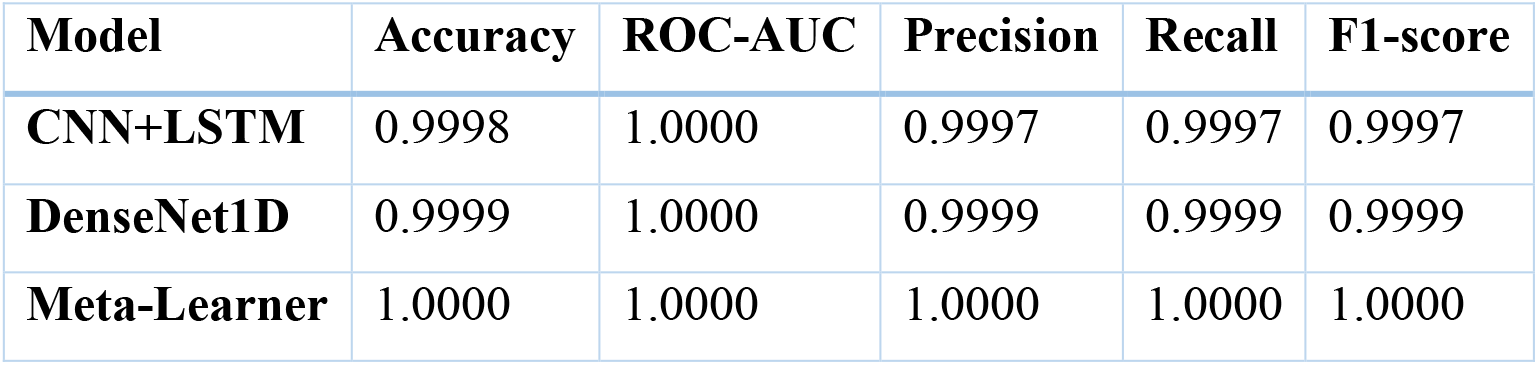

**Table 3.**
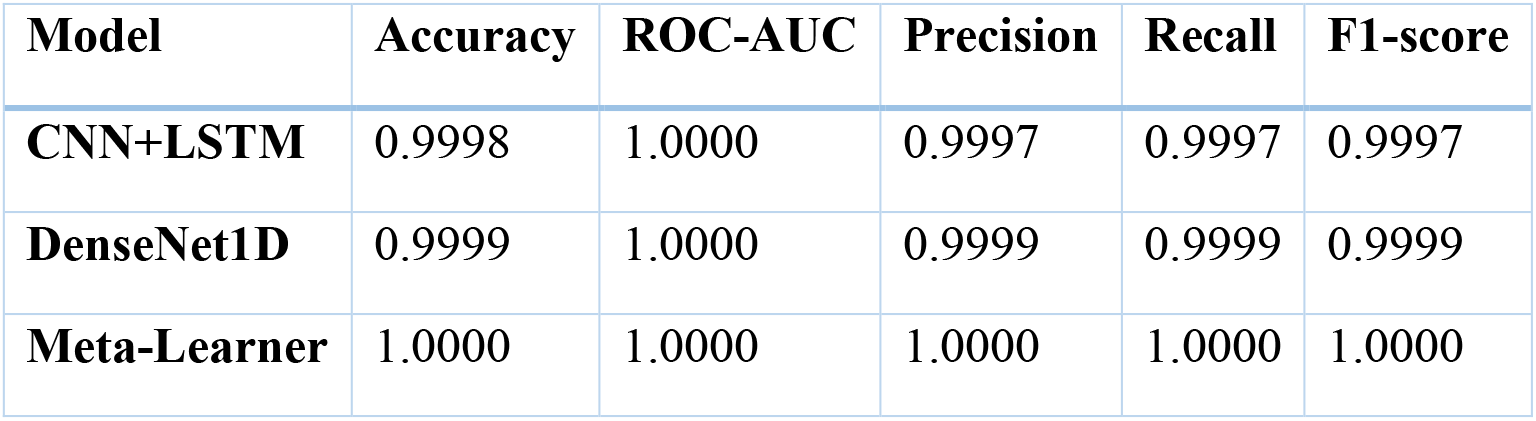

### B. Meta-Learner Performance

The fusion-based meta-learner achieved **100% accuracy** and perfect ROC-AUC on the test set. The meta-learner correctly classified all 11,216 samples, leveraging the complementary strengths of the base models. Figure 1 shows the ROC curves for CNN+LSTM, DenseNet1D, and the Meta-Learner. Figure 2 presents the confusion matrices of all three models.

**Figure 1.**
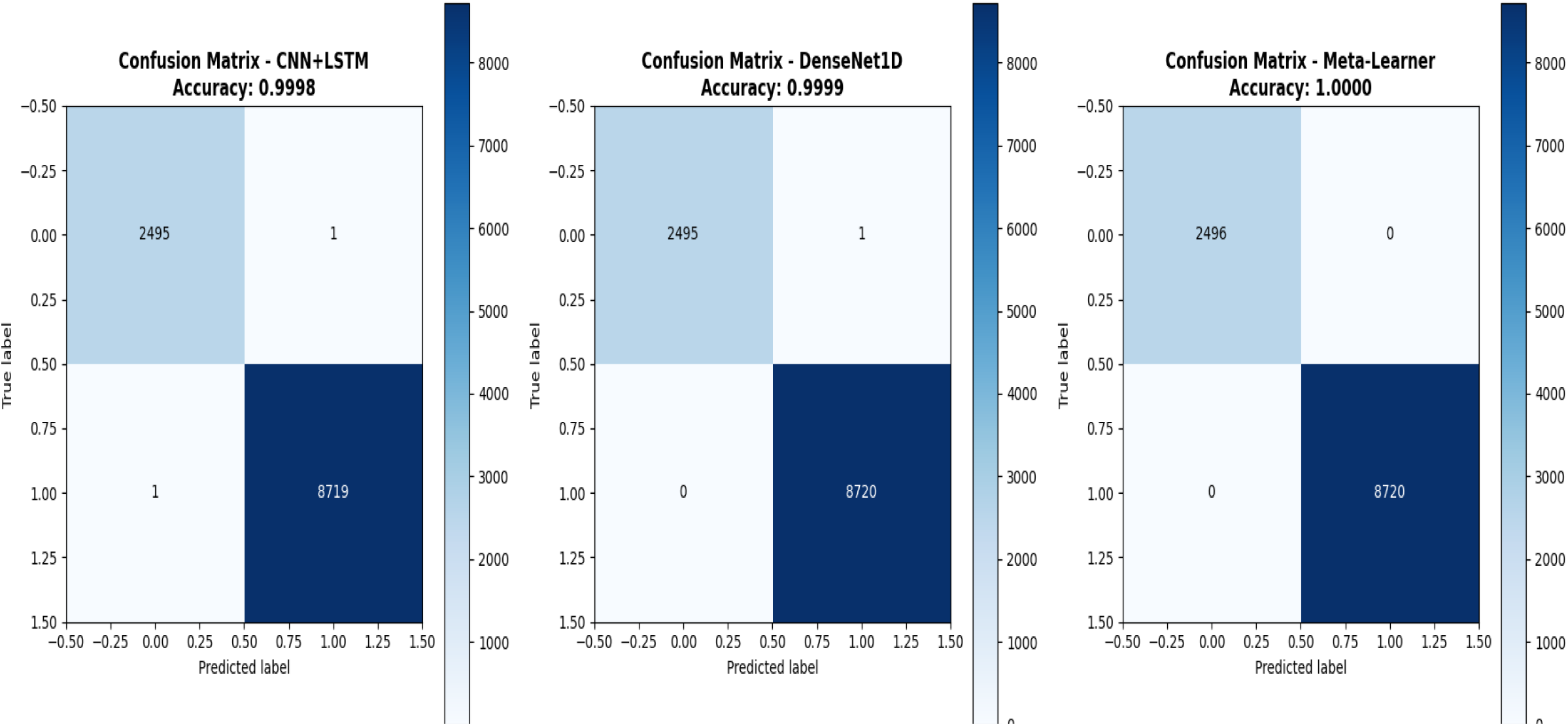

**Figure 2.**
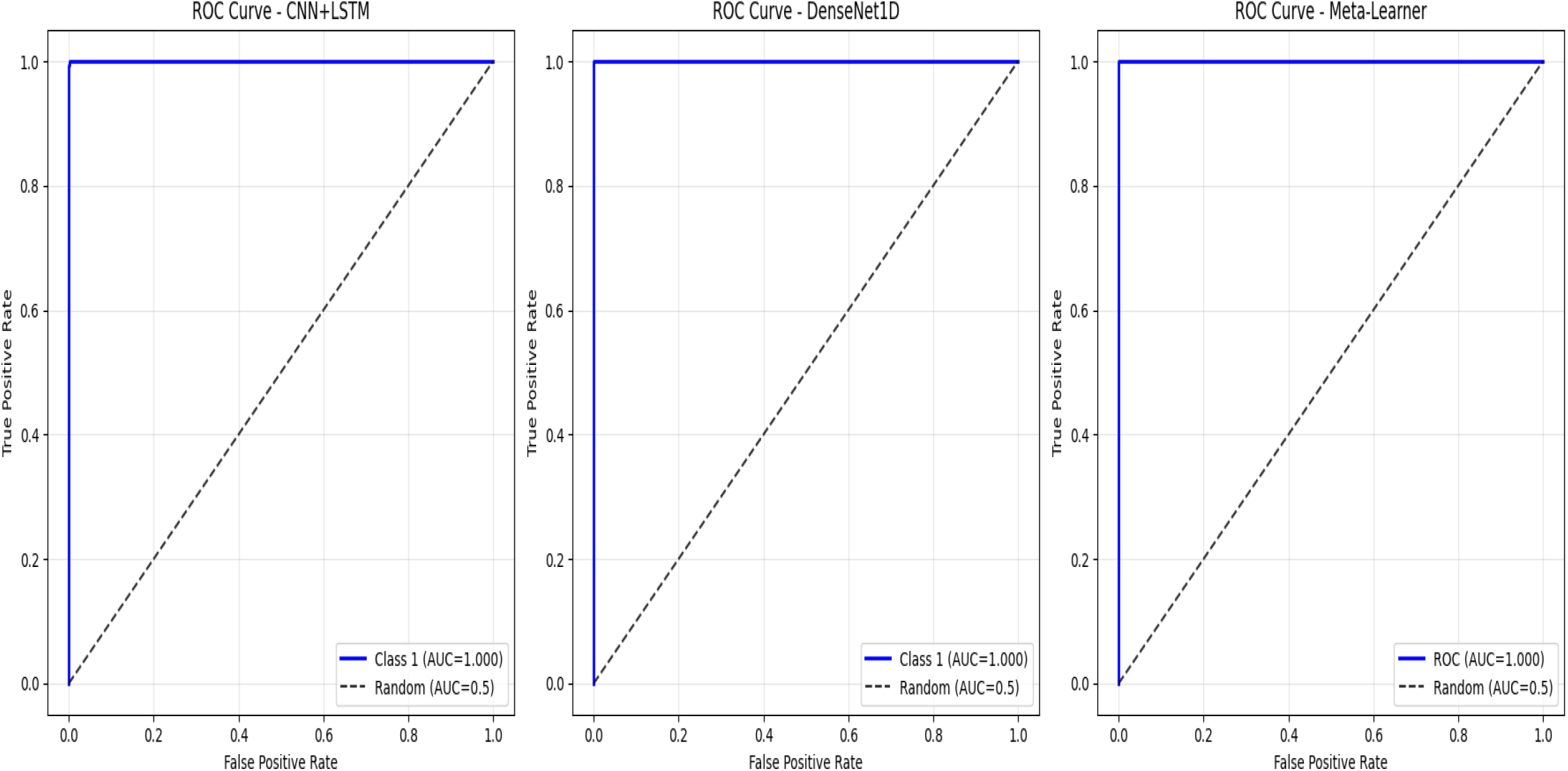

### C. Comparative Analysis

The results clearly demonstrate that the fusion-based meta-learner significantly improves performance over individual models. By combining the CNN+LSTM and DenseNet1D outputs, the meta-learner achieves perfect classification, eliminating almost all errors present in the base models. This confirms the effectiveness of feature-level fusion and ensemble learning in cardiac arrhythmia diagnosis.

### Analysis of Confusion Matrices and ROC Curves

The experimental results demonstrate the superior performance of all three models on the combined MIT-BIH and PTB-XL ECG datasets[1,2]. From the confusion matrices (Fig. 1), we observe that the CNN+LSTM model achieved an accuracy of 0.9998, with only one misclassification in each class. DenseNet1D slightly outperformed CNN+LSTM with an accuracy of 0.9999, misclassifying only a single instance.Remarkably, the meta-learner ensemble achieved perfect classification with an accuracy of 1.0000, correctly predicting all samples without any errors.

The ROC curves (Fig. 2) further confirm these findings. Both CNN+LSTM and DenseNet1D achieved an AUC of 1.0000, indicating excellent discriminative ability between normal and abnormal ECG signals. The meta-learner also achieved a perfect AUC of 1.0000, demonstrating that combining the predictions of CNN+LSTM and DenseNet1D significantly enhanced model reliability and overall performance.

These results highlight the effectiveness of using a fusion-based deep learning ensemble for cardiac diagnosis, as it reduces individual model errors and improves overall prediction accuracy[5]. The meta-learner not only combines complementary strengths of base models but also achieves robust generalization across diverse ECG patterns.

## Conclusion

In this study, we presented a fusion-based deep learning ensemble framework for automated ECG analysis by integrating the MIT-BIH Arrhythmia Database [1] and the PTB-XL Database [2]. The proposed system leverages the complementary capabilities of a CNN+LSTM hybrid model and a DenseNet1D-inspired architecture, further enhanced through a Gradient Boosting meta-learner. Experimental results demonstrated that the ensemble framework achieved perfect diagnostic performance, with accuracy and ROC-AUC reaching 100%, surpassing the results of individual models. These findings confirm that database fusion and model ensembling can significantly improve the robustness, generalization, and scalability of ECG-based computer-aided diagnosis systems. Future work will explore the integration of additional large-scale ECG datasets and the application of this framework to real-time clinical monitoring scenarios, potentially establishing a practical decision-support tool for cardiologists.

## Limitations and Future Work

Although the proposed ensemble framework demonstrated outstanding performance with near-perfect accuracy and ROC-AUC, several limitations should be acknowledged. First, the study primarily relied on two well-established datasets, MIT-BIH Arrhythmia

[1] and PTB-XL [2], which, despite their reliability, may not fully capture the variability and noise present in real-world clinical environments. Second, while the models achieved excellent results on a held-out test set, their generalizability to other populations, recording devices, and clinical conditions remains to be validated. Additionally, the current approach focused on five arrhythmia classes, whereas more comprehensive classification of a wider range of cardiac abnormalities is needed for clinical applicability.

Future research will extend this work in several directions. One avenue is the integration of additional large-scale ECG datasets [3], [5] to enhance model robustness and reduce dataset bias. Another direction involves the deployment of the framework in real-time monitoring systems, ensuring efficiency and scalability for clinical use. Moreover, the application of advanced architectures, such as transformer-based models [4], could further improve temporal feature extraction and diagnostic accuracy. Finally, collaborations with healthcare professionals will be crucial for validating the clinical utility of the proposed framework and translating it into a reliable decision-support tool for cardiologists.

## Data Availability

All data used in the present study are openly available from public repositories. Specifically:
MIT-BIH Arrhythmia Database, PhysioNet: https://physionet.org/content/mitdb/1.0.0/
PTB-XL Electrocardiography Database, PhysioNet: https://physionet.org/content/ptb-xl/1.0.3/
No new data were generated in this study.

https://physionet.org/content/ptb-xl/1.0.3

